# App-Guided Grip Strength Assessment: Feasibility and Validity of Self-administered Testing with a Smart Dynamometer

**DOI:** 10.64898/2026.01.29.26345154

**Authors:** Gabriella Francis, Kayla DeTreux, Mia Enright, Lydia George, Yianna Lambrides, Kartik Mangudi Varadarajan, Winnie Tsui

## Abstract

**Introduction:** Remote grip strength assessment offers potential for scalable at-home health monitoring, yet most validated methods require in-person supervision. The Squegg™ Smart Dynamometer is a Bluetooth-enabled device designed for both supervised and remote app-guided, self-administered testing. Validating self-administered grip strength assessment is essential for clinical and research use. This study evaluated whether self-administered grip strength measurements with the Squegg™ device are comparable to supervised testing, and examined the influence of participant and procedural factors.

**Methods:** In this prospective, within-subject comparative study, 96 healthy adults completed grip strength tests with the Squegg™ device under two modalities: self-administered (app-guided) and supervised (by trained personnel). Covariates included sex, hand dominance, age, education, prior grip testing experience, and test order. Analyses included Shapiro-Wilk tests, ANOVA, and Bland-Altman analysis.

**Results:** Grip strength residuals met normality assumptions. Guidance modality (self-administered vs. supervised) had no significant effect (F_1,270_ = 1.41, p = 0.24). The mean difference between modalities was 0.68 lbs relative to an average grip strength of 83.4 lbs (95% CI: -1.77 to 0.41). Sex explained 45% of between-subject variation. Within subjects, variation was associated with hand dominance (6%), test order (4%), and guidance modality (0.4%). No significant effects were observed for age, education, or prior device experience. Bland-Altman analysis showed consistent agreement across the grip strength range.

**Conclusions:** Self-administered grip strength assessments with the Squegg™ Smart Dynamometer are comparable to supervised testing, supporting its potential for remote patient monitoring. Future work should confirm findings in broader populations, home settings, and longitudinal contexts.

## INTRODUCTION

Grip strength is a widely accepted and essential metric for assessing hand function across a range of clinical conditions. It serves as a key indicator of musculoskeletal health, injury severity, and functional recovery in both adult and pediatric populations^1–3^ Beyond local hand pathology, an extensive body of evidence has established grip strength as a robust biomarker of overall health ^4,5^. Low grip strength has been strongly associated with adverse outcomes such as cardiovascular disease, type 2 diabetes, stroke, cognitive decline, and reduced health-related quality of life, particularly in older adults ^6,7^. In pediatric and adolescent populations, grip strength is similarly linked to cardiometabolic health and bone development ^8–10^. Therefore, accurate grip strength assessment is critical across the lifespan and has relevance in both rehabilitative and preventive health contexts.

Traditionally, grip strength has been measured using hydraulic hand dynamometers such as the Jamar® device. While these tools are considered the clinical gold standard^11,12^, they have limitations including bulkiness, cost, and lack of portability, which has restricted their use to in-clinic settings^13,14^. Moreover, their weight and rigid design pose usability challenges for certain populations—particularly children, older adults, and individuals with pain or joint deformities - potentially impacting measurement accuracy^15^. In recent years, digital hand dynamometers have emerged as promising alternatives^14,16–18^. These devices offer advantages such as lightweight design (e.g. Squegg™= 26g and GripAble™ = 240g, vs. Jamar® = 700g), digital data capture, user-friendly interfaces, and potential for integration with electronic medical records. Critically, they have the potential to enable remote grip strength assessments, which is particularly relevant in the context of telehealth expansion and the growing emphasis on Remote Therapeutic Monitoring (RTM) enabled by recent CPT code changes^19^. Remote assessments can improve access to care for underserved or mobility-limited populations and allow clinicians to monitor progress longitudinally and adjust treatment plans in real time.

However, the shift from in-clinic to home-based grip strength assessment poses several challenges. Remote solutions must be affordable, easy to use without clinical supervision, and capable of delivering reliable, clinically meaningful data. While several digital grip assessment tools exist, none to the best of our knowledge, have been validated for self-administered, at-home use. Thus, the feasibility of remote grip strength testing without therapist oversight remains largely unaddressed in the literature.

One promising tool, the Squegg^TM^ Smart Dynamometer and Hand Grip Trainer **(Fig. 1)**, offers potential to address these gaps. The Squegg™ device is compact (26g), wireless (Bluetooth), and features a soft silicone shell, making it more highly portable and comfortable compared to bulky/rigid traditional dynamometers (>600g). The device pairs with mobile applications—Squegg Core for patients and Squegg PRO for clinicians—that provide standardized grip testing protocols and gamified exercise programs. In previous studies, the Squegg™ has demonstrated concurrent validity and test-retest reliability comparable to standard dynamometers such as the Jamar® and adapted sphygmomanometers, in adult and pediatric cohorts, respectively^13,16,20,21^. Age- and sex-stratified normative values for Squegg™ measured grip strength have also shown trends consistent with those reported for traditional tools, further supporting its use in clinical settings^22^. Despite promising in-clinic validation, limited data exists on the reliability of purely self-administered, app-guided grip strength testing using the Squegg™ system. One study employed Squegg™ in a post-COVID home care kit, where 20 participants used the device over a 30-day recovery period. Results showed significant improvements in grip strength (from 65.0 lbs to 78.2 lbs), aligning with functional metrics such as Timed Up and Go and Sit-to-Stand tests ^23^. However, these assessments were either video-supervised or conducted during home visits (confirmed via direct communication with study authors), leaving the feasibility of completely self-administered, app-guided use unanswered. With respect to other solutions, a systematic review by Heslop et al. sought to evaluate the comparability of remotely collected grip strength data with in-clinic measurements^24^. However, none of the four included studies addressed unsupervised / self-administered use of the same device in both settings limiting the interpretability of findings. Two studies used a different device for at-home (grip-ball/ Nintendo Wii) vs. face-to-face measurements (Jamar dynamometer) ^25,26^; a third study compared video-based supervision to face-to-face assessment using the same device ^27^; while the fourth study compared an unstated device used in-clinic to an app-based grip strength (iFit) assessment ^28^.

**Figure 1.**
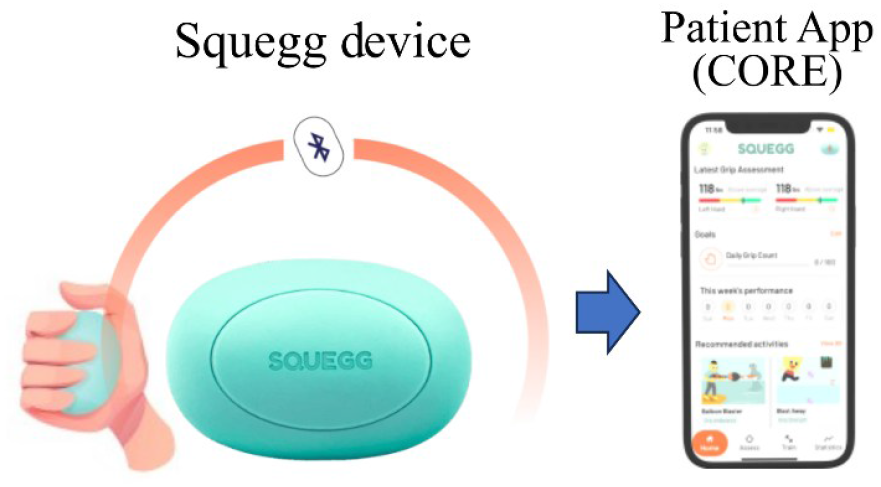
Squegg^TM^ Smart Hand Grip Trainer and companion CORE mobile application

In clinical practice, therapists typically provide verbal encouragement and ensure correct patient posture during grip strength testing through cueing - both factors known to influence test performance ^29–32^. For remote assessments to be valid, these elements must be replicated through technology. The Squegg™ application follows the American Society of Hand Therapists (ASHT) protocol ^12^, and includes visual instructions for guiding posture and positioning, and provides automated prompts for test initiation, holding, swapping of hands, and termination of the grip **(Fig. 2)**. Maximal grip strength is automatically calculated as an average of three trials. Whether this level of automated guidance is sufficient to match the accuracy of supervised assessment remains to be determined, and forms the motivation for this study.

**Figure 2.**
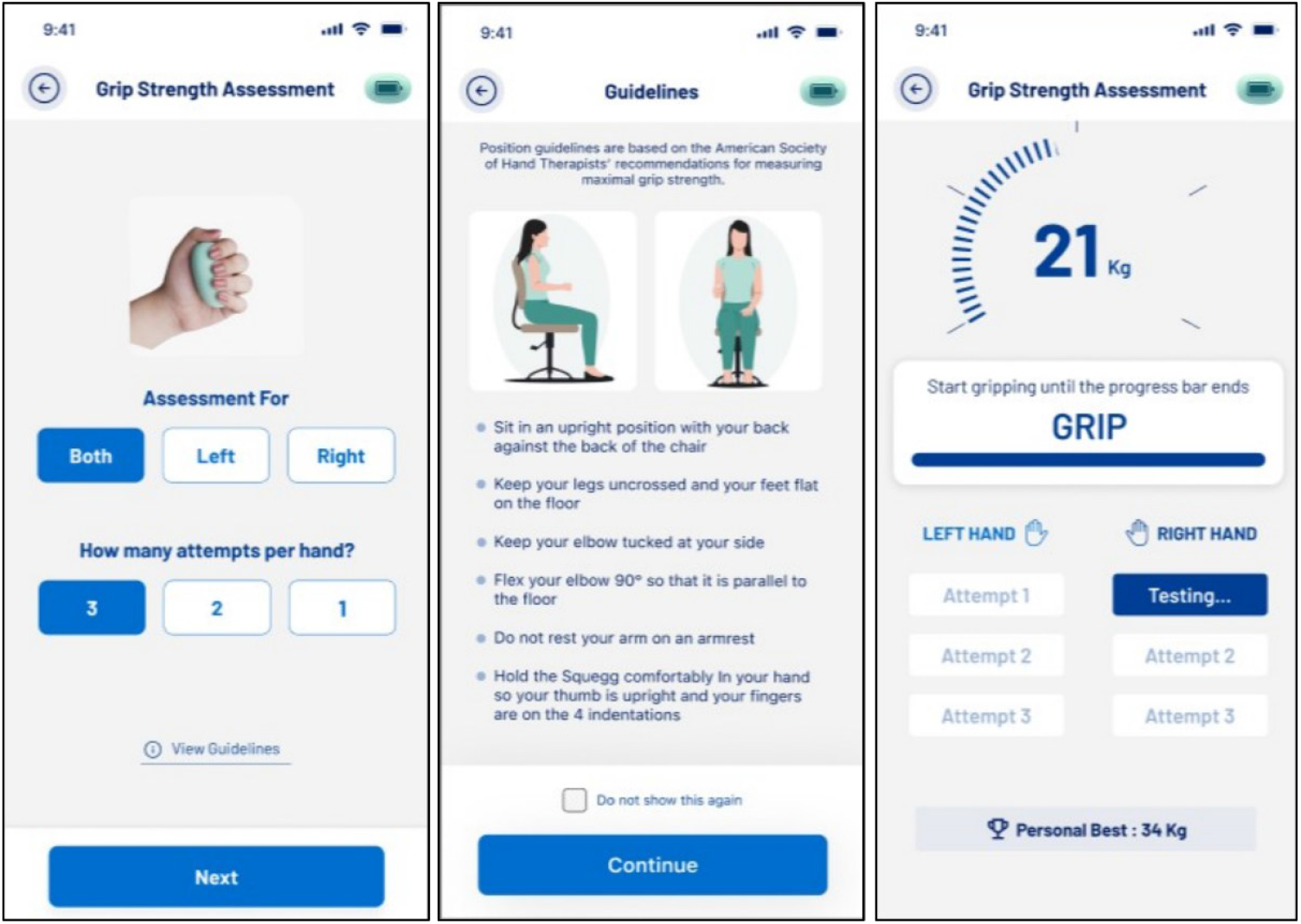
Sample grip strength assessment screens within the Squegg^TM^ Core mobile application

The primary objective of this study was to evaluate the feasibility and reliability of self-administered, app-guided grip strength assessment using the Squegg™ system. Specifically, we assessed the level of agreement between in-clinic, therapist-supervised measurements and self-administered, app-guided measurements among healthy adult participants. Establishing such agreement would support the use of Squegg™ for remote grip strength monitoring, enhancing access to hand therapy services beyond the clinic.

## METHODS

### Hypothesis

The null hypothesis for the study was that there is no statistically significant difference between grip strength measurements obtained through app-guided self-administration, and supervised administration using the Squegg™ Smart Dynamometer.

### Research Design

This study employed a within-subjects experimental design in a prospective healthy cohort who completed grip strength measurements using the Squegg™ Smart Dynamometer under two guidance modalities: app-guided self-administration and supervised administration by trained individuals. To minimize potential influence of natural variations in individuals grip strength over time, the second testing session was conducted within a seven-day period. To mitigate potential learning effect from one session to the next, and to account for order effects, participants were randomly assigned to one of two sequence groups: self-administered first or supervised first. Demographic data, including age, sex, dominant hand, educational attainment, and prior experience with grip strength evaluation, were collected to explore potential covariates.

Participants were recruited based on the following inclusion criteria: a) Age 18 years or older; b) proficiency in English; c) absence of neuromuscular or orthopedic dysfunctions that would impede the performance of maximal grip strength exertions; and d) ability to generate a minimum of 11 pounds of grip force per hand. Exclusion criteria included: a) Mini-Mental State Examination (MMSE) scores below 24, b) uncorrectable visual impairments that would interfere with device usage; c) grip strength measurements below 11 pounds; d) inability to provide informed written consent; and e) Reading comprehension below an 8th-grade level.

### Instruments and Measurements

- **Squegg™ Smart Dynamometer and Hand Grip Trainer** (Squegg Inc., Penbroke Pines, FL, USA): A clinically validated Bluetooth-enabled device (model 1) was the instrument used for grip strength measurement **(Fig. 1)**, paired with its companion patient facing mobile application (Squegg Core version 3.4.0, **Fig. 2)**.
- **Demographic Questionnaire:** A Google Forms questionnaire was used to collect participant demographic information, including age, sex, hand dominance, vision status, educational attainment, prior upper extremity injuries, and prior experience with grip strength testing.
- **Mini-Mental State Examination (MMSE):** This cognitive screening tool was administered to assess cognitive function, with scores below 24 indicating potential cognitive impairment and serving as an exclusion criterion.

### Procedure

The participants were recruited between April to May 2024 through convenient and snowball sampling with Institute Review Board approval. After obtaining informed consent, filling out the demographic survey, and taking the MMSE, each participant was randomly assigned to two groups. Even-numbered participants completed the supervised trial first, followed by the self-administered trial, while odd-numbered participants completed the self-administered trial first, followed by the supervised trial. The ASHT protocol, which calls for participants to sit upright with their elbows flexed at a 90° angle, shoulders adducted, forearm in a neutral position, and feet flat on the ground, was followed. The instructions in the Squegg™ app, and corresponding workflow follow the same ASHT protocol^12^. The average grip strength per hand was automatically calculated from three maximum hand grip strength trials, with participants switching hands between trials. During self-administered use participants followed the written and visual cues provided in the Squegg™ app, which included a visual force meter showing grip as bio-feedback in real time, a load bar to indicate duration of the maximal effort grip squeeze per trial and encouraging prompts for when to squeeze and release. The self-administered testing was done in a private environment to replicate at-home circumstances, with the user left alone to perform assessment. Supervised testing was carried out by investigators (3^rd^ year occupational therapy students KD, ME, LG, YL trained by senior hand/occupational therapists GF, WT) who ensured correct form via verbal and tactile cues, and provided verbal encouragement. Participants were allowed to see the app screen like the self-administered group. For analysis, the grip strength values of all trials of each participant—right-hand supervised, left-hand supervised, right-hand self-administered, and left-hand self-administered —were saved in Google Sheets.

### Analysis of Data

Statistical analysis was conducted using R statistical software, V 4.3 (R Core Team, 2021). Analysis of variance (ANOVA) was used to compare grip strength for self-administered vs. supervised modalities. Grip strength residuals were tested for normality using Shapiro-Wilk tests (Shapiro & Wilk 1965) for both female and male subjects. Initially six covariates were screened for their potential relationship with grip strength – hand dominance (dominant vs. non-dominant hand), sex, age class (18-34, 35-54, or 55+), education level (high school or less, some college, bachelor’s degree or above), test order (i.e. supervised first or self-administered first), and prior experience with grip strength devices (yes or no). Interactions between these six covariates and guidance modality were also tested; for example, supervised measurements could be comparatively higher with the dominant hand but not the non-dominant hand, or self-administered measurements could be lower with older subjects but not younger ones. Based on these initial analyses, main effects or interactions that approached statistical significance (p < 0.10) were included in the final ANOVA used for estimating the effect of guidance modality. In each of these ANOVAs, we incorporated subject as a random effect and tested the effects of guidance modality, hand dominance, and test order against within-subject error. Sex, age class, and prior experience were tested against among-subject error.

In addition, a Bland-Altman plot (Bland & Altman 1986) was constructed to examine the difference between self-administered and supervised measures of grip strength across the range of observed grip strength values. This visual method is useful for identifying discrepancies between the two approaches that would not be apparent when comparing mean values.

## RESULTS

In total 98 participants were recruited, with 2 participants not completing the study. The final study sample comprised 96 participants: 41 males (42.71%) and 55 females (57.29%). The age distribution was as follows: 18-24 years (n=35), 25-34 years (n=32), 35-44 years (n=3), 45-54 years (n=17), 55-64 years (n=9), and 65+ years (n=1). Participants were predominantly right-handed (n=82, 85.42%), with 10 left-handed (10.42%) and 4 ambidextrous (4.17%) participants – these 4 were excluded in any analyses involving hand dominance. Educational attainment varied, encompassing a range from no high school diploma to doctoral degrees (some high school = 3; high school grad = 21, college/trade = 74). While, 25 participants reported prior upper extremity injuries, it did not preclude their ability to perform repeated maximal grip strength tests. A total of 22 participants had previous experience with dynamometry, while 74 participants reported not having any experience. The majority of participants were primary English speakers (N= 92), while four participants reported speaking additional languages (Malayalam, Telugu, Spanish, and Greek).

### Residuals and Covariate Screening

The residuals for grip strength measurements adhered to normality assumptions, with Shapiro-Wilk tests showing non-significant p-values for both females (W = 0.989, p = 0.09) and males (W = 0.988, p = 0.24). Screening of covariates revealed significant main effects for sex (F_1,93_ = 82.4, p < 0.001), hand dominance (F_1,180_ = 28.1, p < 0.001), and test order (F_1,93_ = 7.1, p = 0.009). No significant effects were observed for age group (F_2,92_ = 1.66, p = 0.20), education level (F_2,92_ = 2.00, p= 0.14), or prior experience with grip strength devices (F_1,93_ = 0.05, p = 0.83). Interactions between covariates and guidance modality were non-significant (p > 0.25) and were excluded from the final model.

### Final Model Results

The final ANOVA revealed no significant effect of guidance modality (self-administered or supervised) on grip strength (F_1,270_ = 1.41, p = 0.24). After adjusting for covariates (sex, hand dominance, test order, and a random effect for subject), the mean grip strength measured via self-administration was found to be 0.68 lbs lower than supervised measurements, with 95% confidence intervals (-1.77 to 0.41 lbs) overlapping zero. The magnitude of this difference was small compared to other covariates, such as test order (1.88 lbs), hand dominance (2.44 lbs), and sex (32.3 lbs) **(Fig. 3)**. Based on the analysis of variance, 45% of the between-subject variation was associated with participants’ sex. Within subjects, 6% of the variation was associated with hand dominance, 4% was related to test order, and just 0.4% was explained by guidance modality (self-administered or supervised).

**Figure 3:**
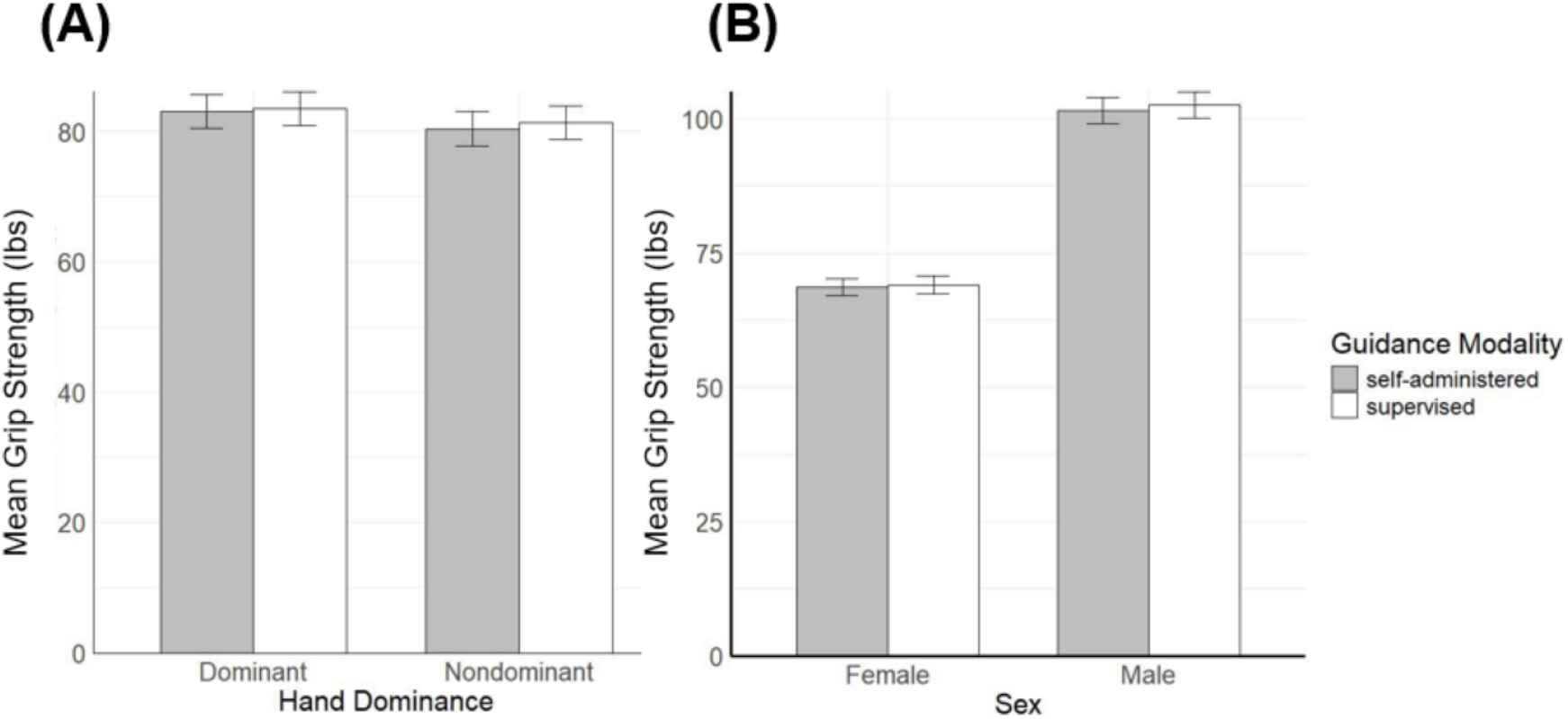
**(A)** Self-administered vs. supervised mean grip strength values for dominant and non-dominant hands of participants. **(B)** Self-administered vs. supervised mean grip strength values for male and female participants. Error bars show one standard error.

### Bland-Altman Analysis

The Bland-Altman plot revealed that differences between self-administered and supervised measurements were consistent across the range of observed grip strength values **(Fig. 4)**. However, participants with higher grip strengths tended to show slightly higher values during supervised testing.

**Figure 4:**
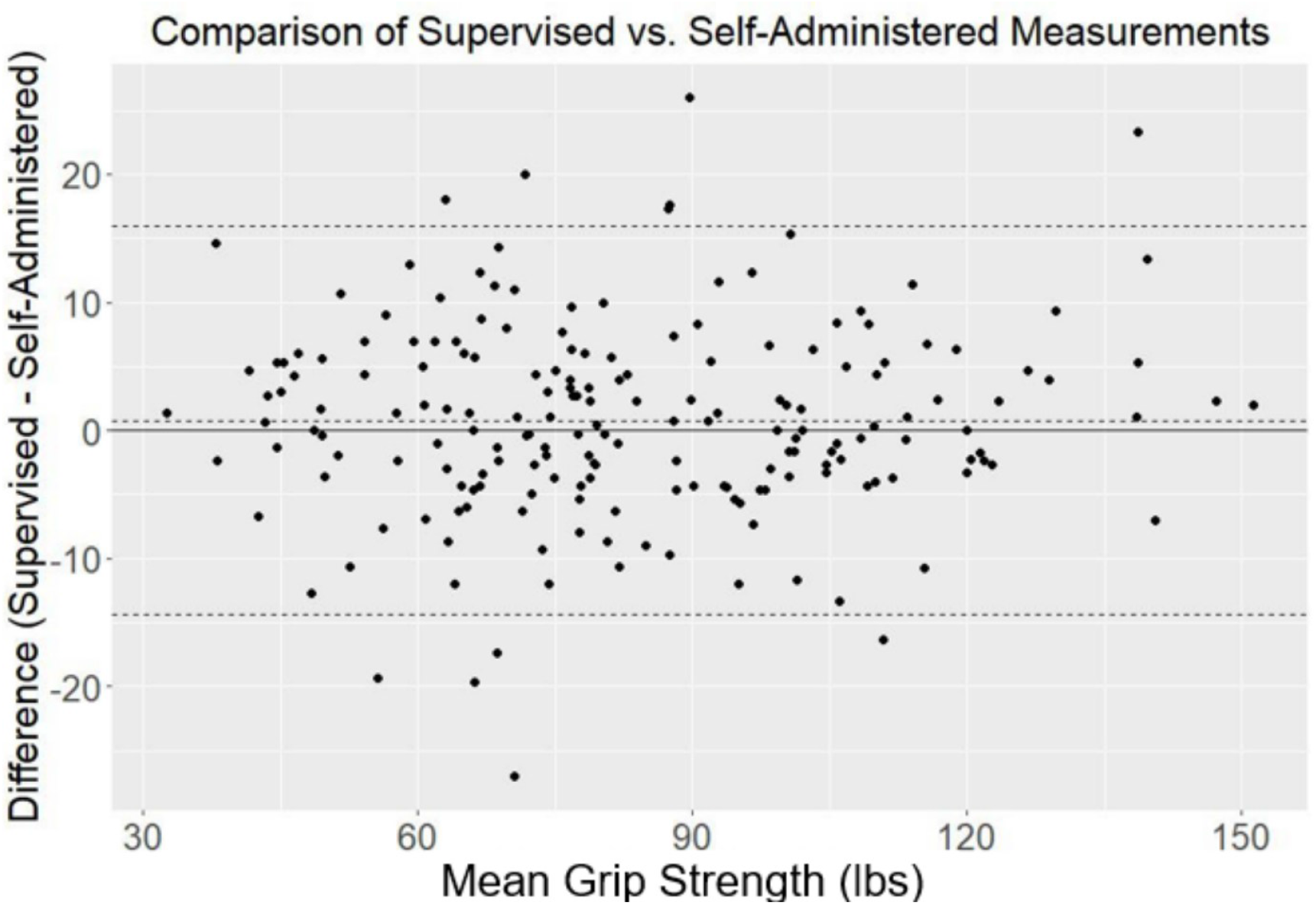
Bland-Altman plot showing the difference between values for supervised vs. self-administered grip strength (lbs; y-axis) as a function of mean grip strength (x-axis). Values above the line correspond to higher values measured under supervised testing and values below the line correspond to higher values measured during self-administered testing.

## DISCUSSION

This study aimed to evaluate the feasibility and validity of self-administered, app-guided grip strength assessments using the Bluetooth-enabled Squegg™ smart hand dynamometer. By comparing self-measured values to those obtained under human supervision, we sought to determine whether purely app-guided assessments could offer a reliable alternative to clinician supervised evaluations—an important consideration for scalable remote health monitoring. The findings support the null hypothesis, showing no statistically significant difference between self-administered and supervised grip strength measurements.

Initial covariate analysis indicated that participant age, education level, and prior experience with grip assessment devices did not significantly influence grip strength outcomes. A subsequent ANOVA confirmed that grip strength measurements did not significantly differ between self-administered and supervised. Instead, most of the observed variance was attributed to sex, hand dominance, and test order. The difference between self-administered and supervised grip strength values was minimal (mean difference = 0.68 lbs, with confidence interval spanning zero), especially when compared to other covariates: test order (1.88 lbs), hand dominance (2.44 lbs), and sex (33.2 lbs). These results affirm the measurement reliability of the Squegg™ device across both testing environments.

The influence of sex and hand dominance on grip strength aligns well with established literature. The ∼3% higher grip strength observed for dominant vs. non-dominant hands of study participants is consistent, though slightly smaller than, the ∼4–7% differences reported previously using Squegg™ and other dynamometers (e.g., Jamar and Takei)^22,33–35^. Similarly, the ∼33% higher grip strength observed in males versus females corresponds with existing reports, including 26% for Squegg™ reported by Varadarajan et al^22^. and ∼38% for Jamar reported by Wang et al^35^.

Interestingly, no significant effect of age group on grip strength was observed in our study. Although this appears to contradict the well-documented decline in grip strength with age^22,35–37^, a closer inspection reveals that the present study sample skewed towards younger age group, with 73% of participants being between 18–44 years of age. Prior studies show that grip strength remains relatively stable in this age range, with noticeable declines typically emerging after age 45^22,35–37^. Therefore, the lack of age-related effect is likely a function of the sample’s age distribution.

Test order showed a modest influence on grip strength, with slightly higher measurements (1.88 lbs) observed for the second test compared to the first. This effect was smaller than that of sex or hand dominance, and may reflect a subtle learning effect resulting in stronger grip strength readings during the second round of testing irrespective of guidance modality (self-administered or supervised). Neither education level nor prior experience with dynamometers had a significant impact on grip strength results. This suggests that the app-guided and verbal instructions provided were intuitive and easily followed, even by those with no prior exposure. Some studies have noted correlations between education level and grip strength, especially among older populations, with higher education linked to positive health behaviors, socioeconomic status, and physical fitness.^38–41^ Nonetheless, prior literature does not point to any impact of educational level on ability to follow verbal instructions during the grip strength assessment.

Bland-Altman analysis revealed consistent agreement between self-administered and supervised grip strength measurements across the full range of values. While participants with higher grip strength showed a slight tendency toward higher supervised scores—possibly due to performance bias or motivation in the presence of an investigator—this effect was minimal and did not compromise the overall comparability of the two testing methods.

Despite promising results of this study, several limitations warrant consideration. Firstly, with 71% of participants being in age-group 18-44, the study sample study skewed towards younger participants. Variation in technological proficiency with age could restrict the generalizability of the study findings. Therefore, future research should prioritize diverse sampling, including older adults and varying tech proficiencies. Nonetheless, given the rapidly growing adoption of technology by older individuals (∼76% of adults 65+ today own a smartphone)^42,43^, we can be optimistic about the feasibility of at-home grip assessments for adults of all ages, perhaps with assistance of caregivers in some cases. Second, although the “remote” assessments were conducted in isolated environments, they did not occur in participants’ actual homes. Environmental differences (e.g., distractions, posture, comfort) in a true home setting could influence performance. Future studies should aim to evaluate remote testing in natural home environments to improve ecological validity. Third, assessments were conducted within a short follow-up period (within seven days) to avoid natural grip strength variation over time; however, this could introduce learning effects (learning from first session impacting the second session). Randomization of testing order with one group performing self-administered assessment prior to supervised assessment, and the other group performing them in opposite order helped to mitigate this concern. Nonetheless, in future studies impact of a longer interval between testing sessions could be explored. Fourth, all participants were fluent English speakers and had completed at least a high school education. This limits the generalizability of findings to populations with lower literacy levels or limited English proficiency. Although the Squegg™ app currently supports several languages (English, Spanish, German, and French), broader linguistic and cultural validation is necessary to ensure equitable usability. Finally, this study focused on single-time-point assessment. Long-term use of the device at-home could raise additional considerations around patient compliance and engagement. User motivation and engagement may wane over time—a common challenge in mobile health (mHealth) tools. Prior research on mHealth app usage has identified boredom, declining motivation, and loss of interest in personal health goals as key reasons for abandonment^44^. Future longitudinal studies should assess whether grip strength measurements remain accurate over time in the absence of continued supervision and whether users remain engaged in regular self-assessment. Such work could inform the development of app-based features (e.g., gamification, reminders, feedback) to enhance long-term adherence.

Overall, this study offers compelling early support for the Squegg™ Smart Dynamometer as a viable solution for remote grip strength assessments. The minimal differences between supervised and self-administered measurements suggest that the device can serve as a reliable tool for longitudinal monitoring, particularly in rehabilitation, aging research, and chronic condition management. For clinicians, this introduces the possibility of expanding assessment reach beyond clinic walls, allowing for frequent, self-administered tracking of hand function at home. This could reduce the need for in-person appointments, lower costs, and improve adherence to therapeutic interventions. These findings also have important implications for expanding access to care, particularly for individuals in rural or underserved areas who may face geographic or transportation barriers. By enabling accurate remote assessment, this approach promotes greater health equity and supports more inclusive, continuous care regardless of a patient’s location. For researchers, the validation of a remote-capable device opens opportunities for large-scale data collection in population health and epidemiological studies without geographic constraints.

## CONCLUSION

In conclusion, this study demonstrates the feasibility and validity of self-administered grip strength measurements using the Squegg™ Smart Dynamometer. Comparable results across supervised and self-administered modalities validate the utility of this technology for remote assessment. While further research is needed to explore its application across more diverse populations and over extended periods, the device holds strong potential as a scalable tool for remote home-based hand function monitoring.

## Data Availability

All data produced in the present study are available upon reasonable request to the authors.

## DECLARATIONS

### Ethical approval

Ethical approval for this study was obtained from Long Island University Institutional Review Board (Protocol ID: 24/02-017).

### Informed consent

Written informed consent was obtained from all subjects before the study.

## Acknowledgements

We sincerely thank Prof. David M. Marsh (Professor of Biology, Washington and Lee University, Lexington, VA) for conducting statistical analysis for this study.

